# Is delirium after stroke associated with dysregulation of Hypothalamic pituitary axis?

**DOI:** 10.1101/2024.03.11.24304087

**Authors:** AJ Barugh, AMJ MacLullich, SD Shenkin, M Allerhand, GE Mead

## Abstract

Delirium after stroke is a serious condition associated with poorer longer term cognition. However the mechanism of delirium is poorly understood. The aberrant stress response has been postulated as a mechanism for delirium.

**Aim:** To explore the relationship between cortisol dysregulation and delirium over the first year after stroke in a prospective cohort study of patients admitted to an acute stroke unit.

**Methods:** Consecutive patients admitted to an acute stroke unit over a one year period were identified and recruited if they were aged 65 or over and not taking steroids. Patients with incapacity were included if proxy consent could be obtained. Baseline data included stroke severity, cognition, illness severity, and pre-stroke cognition. Patients were assessed at 1, 3, 5, 7, 14, 21, 28 days, 4 months and 12 months for delirium. Salivary samples were taken morning and evening for cortisol analysis.

**Results:** Of the 831 patients screened, 304 met inclusion criteria and of these 95 agreed to participant. Twenty-six (27%) had delirium at some point during the 12 months follow-up. Delirium was associated with increasing age (mean age 83.5 years vs 74 years, p<0.001), being female (62% vs 23%, p=0.013), not independent in pre-stroke activities of daily living (35% vs 33%), higher IQCODE score median 3.56 vs 3.19), worse stroke severity (median National Institute of Stroke Scale 5 vs 8.5) p=0.009) and having had a total anterior circulation stroke (p<0.001). Univariable analyses identified several associations between delirium and cortisol in the first 28 days but not at 4 or 12 months. However, on multivariable analyses there were no significant associations between delirium and cortisol at any time point e.g. odds ratio for median 9am cortisol 0.95 (95% CI 0.89 to 1.01, p=0.08).

**Conclusion:** There was no independent association between delirium and cortisol dysregulation after stroke. If an association does exist, it is likely to be small.

## Background

Delirium is common after stroke; a 2019 systematic review reported a frequency of between 1.4% to 75.6% of patients [1]. A subsequent 2019 study of over 700 patients reported that about a quarter developed delirium in an acute setting [2]. Delirium is independently associated with poorer functional outcomes [3] and needing 24 hour care [4]. Delirium in the older population predicts later dementia [5].

The underlying mechanisms of post-stroke delirium are uncertain. Hormonal abnormalities, including dysregulation of the hypothalamic-pituitary-adrenal (HPA) axis, have long been considered to be potentially important in its aetiology [6,7]. After hip fracture, high cerebrospinal fluid cortisol is associated with delirium. Treating patients in the intensive care unit setting with steroids is associated with longer term cognitive impairment [6]. If HPA axis dysregulation (e.g. impaired negative feedback regulation of cortisol, loss of the circadian rhythm), is associated with delirium after stroke, and if this relationship is causal, then this could provide a target for the prevention and/or treatment of post-stroke delirium; and this might reduce the longer term risk of cognitive impairment and dementia after stroke.

Stroke is associated with a stress response and raised cortisol [8]. There is an association between higher bedtime cortisol after ischaemic stroke and worse longer term cognition [9], but it is not known whether delirium mediates this relationship. There are a few studies exploring the relationship between cortisol and delirium after stroke; but these were small, not all potential confounders were corrected for, and some measured cortisol at a single time point after stroke [10–13].

Therefore, the aim of this study is to test the hypothesis that delirium after stroke is associated with higher circulating levels of cortisol, not just at a single time point after stroke, but at multiple time points in the first year after stroke.

## Methods

This was a prospective cohort study of patients admitted to an acute stroke unit in a teaching hospital from October 2012 for one year and followed up for a year. Inclusion criteria were clinically confirmed stroke, age ≥ 60 years and onset within previous 120 hours (or time since last seen well for those occurring overnight) at the time of consent. Exclusion criteria were transient ischaemic attack, subarachnoid haemorrhage, current or recent use (within 6 months) of oral or inhaled corticosteroids, active alcohol withdrawal and inability to speak English prior to stroke. A clinical research fellow (AJB) approached potentially eligible patients; those with capacity consented for themselves, proxy consent was obtained from those without capacity. Easy access information materials were available for people with aphasia.

### Baseline clinical data

AJB collected the following data at time of recruitment: pathological stroke type (ischaemic or haemorrhagic), Oxfordshire Community Stroke Project classification (OCSP) and National Institute of Health Stroke Scale score (NIHSS). The National Adult Reading Test (NART) [14] and The Informant Questionnaire on Cognitive Decline in the Elderly (IQCODE) [15] were used to assess premorbid cognition. Heart rate, blood pressure, oxygen saturations and temperature, medications and admission blood results were recorded at the time of recruitment. The APACHE II (The Acute Physiology and Chronic Health Evaluation System II [16] was used as a measure of current illness burden. Past medical history was extracted from case notes.

### Delirium assessments

Participants were assessed for delirium after the onset of stroke at 1, 3, 5, 7, 14, 21, 28 days, 4 months and 12 months. Those recruited after day 1 were seen from day 3 or day 5 onwards. Those discharged from hospital after a short admission were seen as per protocol whilst an in-patient and were then seen at home on day 28 and at 4 months and 12 months. AJB had been trained in screening and diagnosing delirium by two experienced raters.

The Confusion Assessment Method ICU [17] which includes the Richmond Agitation and Sedation Scale (RASS) [18], and the Delirium Rating Scale-Revised-98 [19] were used to identify delirium and its severity. The OSLA was used to assess alertness [20]. The Chart Confusion Assessment Method [21] was also performed. Attention, a core feature of delirium, was assessed by digit span (forwards and backwards). A validated computerised test for attentional deficits in delirium [22] was used for all participants except for those with a visual field deficit. A diagnosis of delirium was made based on the four DSM-IV criteria (acute onset or fluctuating course, inattention, disorganised thinking and altered level of consciousness).

### Cognition

At baseline, 1 month, 4 months and 12 months, cognition was assessed using the Montreal Cognitive Assessment MoCA [23,24].

At 1 month, 4 month and 12 month follow-up, a detailed neuropsychological battery was also administered; these data will be reported separately.

### Other outcome measures

At each of the follow-up visits (1 month, 4 months and 12 months), the Nottingham Extended Activities of Daily Activity score was performed [25]. We also extracted information from clinically performed brain imaging on lesion location, atrophy and white matter disease which will be reported separately.

### Salivary cortisol sampling

Saliva was collected by the research fellow at recruitment at 0930 and 1530, and at the same times on each subsequent assessment day. For follow-ups (1 month, 4 months and 12 months) participants with intact swallowing who could follow written instructions at hospital discharge were sent saliva swab and instructions for collection on the day of the researcher’s visit. They collected a saliva sample on waking on the morning of the researcher’s visit, stored it in the fridge, and the research fellow collected the second sample at around 1530. For those who were unable to collect their own saliva sample, two visits were made to the participant’s home, one in the morning and one in the afternoon. Samples were collected using Salimetrics Oral Swabs (for those with an intact swallowing mechanism, as judged by the clinical team looking after the participant) or Salimetrics Children’s Swabs (for those with dysphagia), placed in a sealed tube, centrifuged for 10 minutes at 3000rpm, at room temperature, and then stored at −80 °C. The Dresden LabService Gmbh (Tatzberg 47-49, 01307, Dresden, Germany) performed the analysis using Enzyme-Linked Immunosorbent Assays (ELISA).

### Statistical analysis

Analyses were performed using Statistical Package for the Social Sciences (SPSS) version 14 and 19, except for the random effects modelling using the R statistics package.

### Power calculation

Assuming a sample size of 120 participants, and a delirium incidence of 30-40 %, the main analyses were comparisons of cortisol levels at each time point between patients with (∼N=40) and without (∼N=80) delirium. With alpha set at p=0.05, we had >80% power to detect medium-sized (Cohen’s d=0.5) differences.

### Statistical Analysis

Participants were divided into two groups, those who developed delirium at *any time point* and those who did not, and descriptive analyses were performed for the two groups.

Morning (am) and afternoon (pm) cortisol levels from each time point were plotted on simple line graphs. Am:pm ratios were calculated and plotted to explore cortisol variability throughout the day. Histograms to demonstrate the distribution of the data were plotted. A Mann Whitney U test was performed to compare unadjusted cortisol levels between the two groups.

### Bivariable analysis

Spearman’s correlations were used for the relationship between cortisol levels and baseline characteristics including age, NIHSS score, admission APACHE II score, IQCODE and Charlson Comorbidities Index. Biserial correlations were used for cortisol levels and sex (binary outcome, male or female). Spearman’s correlations were used for the relationship between cortisol levels and continuous measures of delirium taken on the same day as the cortisol samples (the DRS-R98 and the Del-box); and biserial correlations were used for the relationship between cortisol and dichotomised (positive or negative) measures of delirium (CAM-ICU (taken on the same day as the cortisol samples) and delirium diagnosis at any time point during the 12 months).

For those who developed delirium, Spearman’s correlations were used to explore the relationship between cortisol levels on the day of delirium diagnosis and delirium severity (as measured by the DRS-R98) on the same day. Finally, biserial correlations were used to investigate the relationship between the peak morning and peak afternoon cortisol levels during the first 14 days after stroke (morning and afternoon samples taken on the same day) and diagnosis of delirium at any point throughout the study period and to explore the relationship between median cortisol in the first 14 days after stroke and delirium diagnosis.

### Multivariable analysis

Multicollinearity was tested using variance inflation factor. Binary logistic regression, using the enter method, was used to explore the relationship between delirium diagnosis at any time throughout the study (as a binary outcome, yes or no) and median salivary cortisol levels in the week after stroke (day 0-7). Models were constructed using median morning and median afternoon cortisol levels. The first week was chosen as cortisol levels were found to be high in the first week after stroke in a systematic review [8]). A second set of models, using binary logistic regression were constructed using the peak cortisol levels (morning, afternoon and the ratio of morning to afternoon) during the first 14 days after stroke. The first 14 days was used for this analysis (rather than the first 7 days) in order to capture any later peaks. Important covariates selected a priori were age, sex, NIHSS score, IQCODE, Charlson Comorbidities index and APACHE II score, thus the analysis was designed to estimate the effect of cortisol on delirium independent of stroke severity, acute illness severity, chronic illness burden and prior cognitive impairment.

Finally a generalised linear random effects model was fitted to investigate the effect of cortisol (morning, afternoon and ratio of morning: afternoon) over time on presence or absence of delirium. The outcome variable was presence of delirium at any point during the study (yes or no), and the predictor variable was salivary cortisol level. The model was specified with binomial errors and a logit link function, which is appropriate for predicting the odds of group membership using a binary outcome variable (in this instance delirium, yes or no). The analysis was conducted using the GLMER function in the LME4 R statistics package.

## Results

Of the 831 patients screened during the recruitment period, 304 met inclusion criteria and of these 95 agreed to participant. Seventy-four participants were followed up at 4 months and 68 were followed up at 12 months (attrition was mainly due to death, ongoing ill health and disability). Twenty-six (27%) had delirium at some point throughout the 12 month study period. Delirium was diagnosed at a median of day 5 after stroke (IQR day 4-day 7); median delirium severity score (DRS-R98) was 22 (IQR 16-26). Delirium was associated with increasing age, being female, less independent in pre-stroke ADLs, pre-existing cognitive impairment (IQCODE score), increasing stroke severity and having had a TACS on univariable analysis (table 1).

**Table 1.**
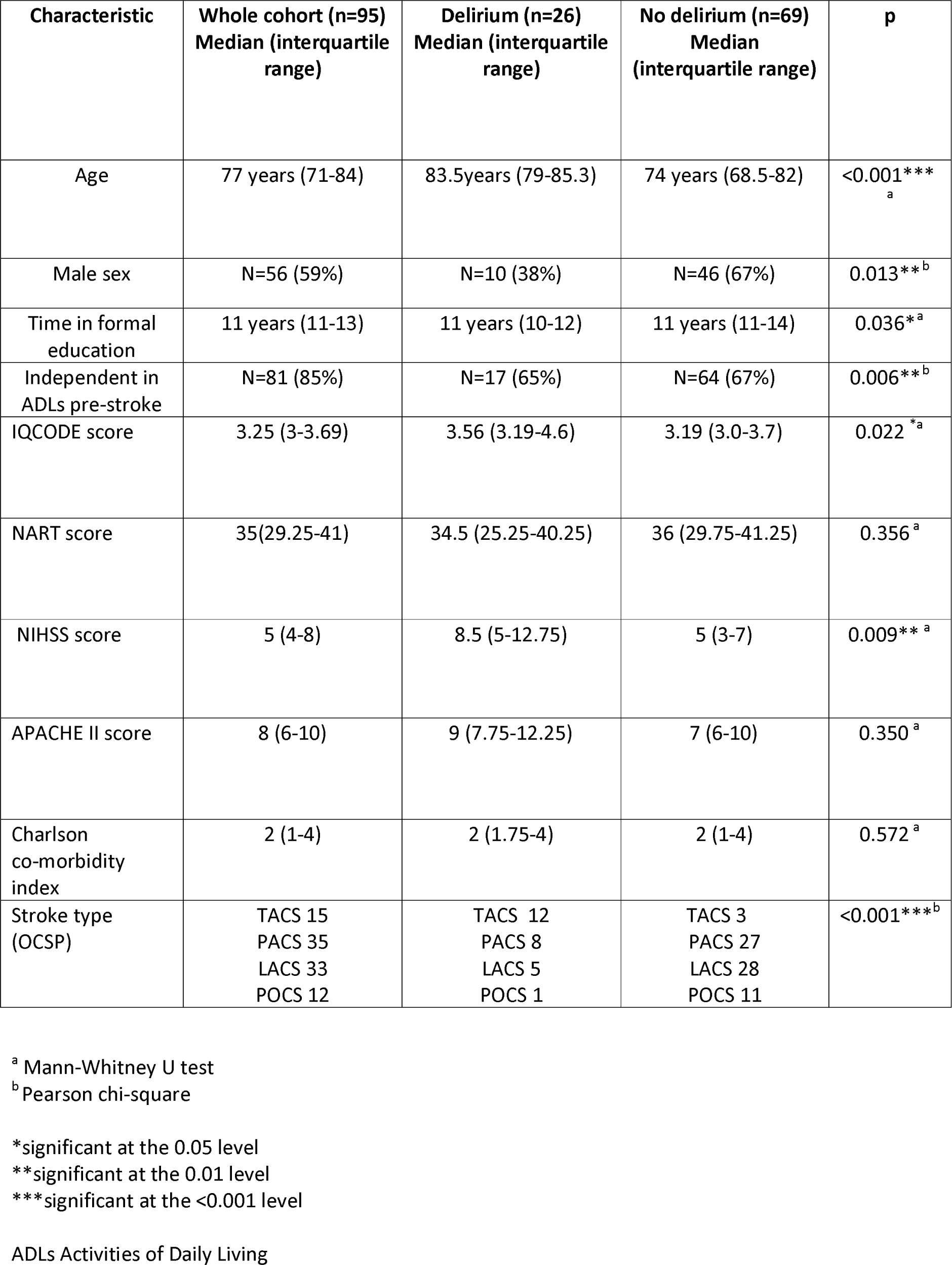
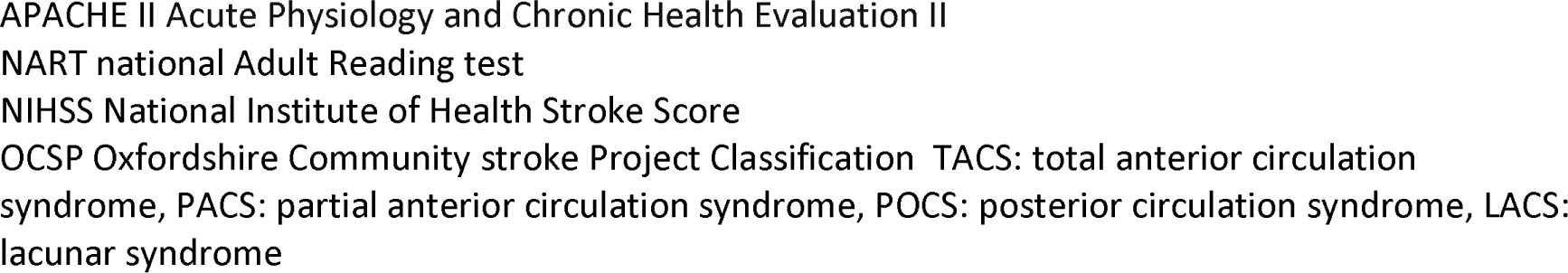
Characteristics of those developing delirium at at least one time point during the study.

The number of patients available for cortisol measurements ranged from 37 (9am cortisol day 14) to 77 (9am cortisol day 28). As expected, cortisol levels were higher in the morning or afternoon at each time point. Descriptive data for cortisol measurements and relationship between those and without delirium are shown in table 2. Of the 21 cortisol measurements shown, there were statistically significant associations (p<0.05) between delirium and seven of these measurements.

**Table 2.** Cortisol levels at each time point compared with those who developed delirium at any time point this those who didn’t.

| Cortisol sample<br>(nmol/L) | No<br>delirium<br>(n=69) |  |  | Delirium<br>(n=26) |  |  | p0 |
| --- | --- | --- | --- | --- | --- | --- | --- |
|  | n | Median | IQR | n | Median | IQR |  |
| 9am cortisol day 3 | 39 | 22.58 | 14.16-30.41 | 9 | 29.76 | 25.10-38.89 | 0.04* |
| 3.30pm cortisol day 3 | 37 | 13.15 | 8.53-18.08 | 8 | 20.14 | 10.83-54.81 | 0.05 |
| Ratio am:pm cortisol day 3 | 36 | 1.72 | 1.26-2.28 | 7 | 1.41 | 1.02-3.03 | 0.55 |
| 9am cortisol day 5 | 49 | 24.33 | 18.5-30.42 | 21 | 24.53 | 18.81-33.20 | 0.74 |
| 3.30pm cortisol day 5 | 48 | 13.45 | 9.3-17.57 | 18 | 18.18 | 13.11-20.95 | 0.04* |
| Ratio am:pm cortisol day 5 | 46 | 1.87 | 1.28-2.58 | 18 | 1.44 | 0.95-1.75 | 0.02* |
| 9am cortisol day 7 | 35 | 21.72 | 16.75-33.06 | 17 | 23.12 | 16.07-27.66 | 0.66 |
| 3.30pm cortisol day 7 | 32 | 11.58 | 9.25-16.73 | 15 | 14.48 | 10.95-16.96 | 0.58 |
| Ratio am:pm cortisol day 7 | 31 | 1.51 | 1.16-2.46 | 14 | 1.57 | 1.40-2.26 | 0.93 |
| 9am cortisol day 14 | 26 | 18.35 | 15.17-24.51 | 11 | 32.18 | 17.15-43.17 | 0.01** |
| 3.30pm cortisol day 14 | 19 | 11.92 | 9.32-15.86 | 11 | 15.94 | 12.80-20.61 | 0.02* |
| Ratio am:pm cortisol day 14 | 18 | 1.65 | 1.24-2.38 | 9 | 1.77 | 1.02-2.21 | 0.94 |
| 9am cortisol day 28 | 59 | 21.92 | 17.88-30.82 | 18 | 31.25 | 16.48-39.06 | 0.22 |
| 3.30pm cortisol day 28 | 60 | 12.19 | 8.95-15.66 | 14 | 16.41 | 11.07-22.54 | 0.03* |
| Ratio am:pm cortisol day 28 | 54 | 1.93 | 1.4-2.93 | 11 | 1.66 | 1.41-2.00 | 0.48 |
| 9am cortisol 4 months | 50 | 22.43 | 15.65-27.03 | 15 | 29.81 | 18.66-34.15 | 0.04* |
| 3.30pm cortisol 4 months | 60 | 12.52 | 7.9-15.72 | 14 | 14.46 | 8.93-19.11 | 0.28 |
| Ratio am:pm cortisol 4 months | 48 | 1.83 | 1.16-2.66 | 11 | 1.98 | 1.41-2.21 | 0.85 |
| 9am cortisol 12 months | 53 | 24.61 | 16.64-37.6 | 15 | 23.57 | 18.2-36.5 | 0.91 |
| 3.30pm cortisol 12 months | 53 | 11.33 | 6.22-22.17 | 14 | 13.94 | 7.94-22.56 | 0.42 |
| Ratio am:pm cortisol<br>12 months | 50 | 2.09 | 1.18-3.4 | 13 | 1.9 | 1.33-3.26 | 0.97 |

Table 3 shows correlations between cortisol levels at each time point and delirium measures at the same time point. Afternoon cortisol levels significantly correlated with the three delirium rating scores (DRS-R98, DEL-box and CAM-ICU) on day 3 and day 5 after stroke and correlated with one score (DRS-R98) on day 14 and with two (DEL-box and CAM-ICU) on day 28. Morning cortisol levels significantly correlated with the three delirium rating scores on day 14 and with the CAM-ICU alone on day 28. The ratio of morning to afternoon cortisol correlated significantly with the CAM-ICU on day 3, with all three delirium rating scores on day 5 and with the DRS-R98 on day 28. There were no significant correlations between any of the cortisol measurements taken at 4 months and 12 months and measures of delirium.

**Table 3.** Correlations between cortisol levels at each time point and measures of delirium taken at the same time point.

| Cortisol sample (nmol/L) | DRS-R98 (Spearman's rho) | Del-box (Spearman's rho) | CAM-ICU (biserial correlation) |
| --- | --- | --- | --- |
| 9am cortisol day 3 | 0.226(p=0.063) | -0.150(p=0.182) | 0.187(p=0.104) |
| 3.30pm cortisol day 3 | 0.322(p=0.015)* | 0.412(p=0.005)** | -0.66(p<0.001)*** |
| Ratio am:pm cortisol day 3 | -0.182(p=0.121) | 0.167(p=0.161) | -0.256(p=0.049)* |
| 9am cortisol day 5 | 0.07(p=0.29) | 0.021(p=0.43) | -0.010(p=0.47) |
| 3.30pm cortisol day 5 | 0.30(p=0.007)** | 0.259(p=0.019)* | -0.239(p=0.03)* |
| Ratio am:pm cortisol day 5 | 0.273(p=0.01)** | 0.336(p=0.004)** | 0.34(p=0.003)** |
| 9am cortisol day 7 | 0.067(p=0.318) | 0.029(p=0.424) | -0.176(p=0.106) |
| 3.30pm cortisol day 7 | 0.126(p=0.200) | -0.344(p=0.012) | -0.006(p=0.483) |
| Ratio am:pm cortisol day 7 | 0.071(p=0.321) | 0.260(p=0.051) | 0.132(p=0.194) |
| 9am cortisol day 14 | 0.343(p=0.02)* | -0.394(0.012)* | 0.453(p=0.003)** |
| 3.30pm cortisol day 14 | 0.369(p=0.025)* | -0.144(0.233) | 0.131(0.249) |
| Ratio am:pm cortisol day 14 | 0.171(p=0.201) | -0.315(p=0.062) | 0.195(p=0.170) |
| 9am cortisol day 28 | -0.135(p=0.121) | -0.175(p=0.076) | 0.234(p=0.022)* |
| 3.30pm cortisol day 28 | 0.167(p=0.077) | -0.37(p=0.001)** | 0.208(p=0.041)* |
| Ratio am:pm cortisol day28 | 0.330(p=0.004)** | 0.209(p=0.059) | -0.104(p=0.210) |
| 9am cortisol 4 months | 0.090(p=0.243) | 0.080(p=0.267) | 0.180(p=0.155) |
| 3.30pm cortisol 4 months | 0.100(p=0.204) | 0.092(p=0.220) | NA |
| Ratio am:pm cortisol 4 months | -0.072(p=0.299) | 0.068(p=0.308) | NA |
| 9am cortisol 12 months | 0.008(p=0.008) | -0.016(p=0.449) | 0.050(p=0.345) |
| 3.30pm cortisol 12 months | 0.205(p=0.050) | -0.086(p=0.246) | 0.168(p=0.089) |
| Ratio am:pm cortisol 12 months | -0.186(p=0.074) | 0.047(p=0.358) | -0.076(0.279) |

\*significant at the 0.05 level
\*\*significant at the 0.01 level
\*\*\*significant at the <0.001 level
NA- Not applicable as CAM-ICU negative for all subjects

The assumptions for logistic regression including linearity and independence of errors were tested and met. Multicollinearity was tested using variance inflation factor statistics. There was no independent association between cortisol and delirium on multivariable analyses (table 4).

**Table 4.** Summary of the Multivariable analysis showing the independent predictors of delirium.

| **Cortisol measure** | **Number of participants** | **Proportion of variance accounted for (%)** | **Statistically significant predictors (p<0.05) of delirium** |
| --- | --- | --- | --- |
| Median 9am cortisol (days 0-7): | 86 | 33-48 | Age<br>NIHSS on admission |
| Median 3.30pm cortisol (days 0-7): | 83 | 27-40 | Age<br>NIHSS on admission |
| Ratio of median morning to afternoon cortisol (days 0-7): | 83 | 30-45 | NIHSS on admission |
| Peak 9am cortisol (days 0-14) | 84 | 37 to 55 | Age<br>NIHSS on admission<br>IQCODE score |
| Peak 3.30pm cortisol (days 0-14) | 82 | 27 to 41 | Age |
| Ratio of peak morning: afternoon cortisol (days 0-14) | 80 | 34 to 52 | Age<br>NIHSS on admission<br>IQCODE<br>APACHE II |

Table 5 shows the univariate association between cortisol and clinical variables.

**Table 5.** Correlations between cortisol levels and baseline characteristics.

| **Cortisol sample (nmol/L)** | **Age (years)** | **NIHSS score on admission** | **APACHE II score on admission** | **IQCODE score** | **Charlson Comorbidities Index score** | **Sex** |
| --- | --- | --- | --- | --- | --- | --- |
| 9am cortisol day 3 | 0.357 (p=0.013)* | 0.425(p=0.003)** | 0.503 (p=<0.001)*** | 0.041 (p=0.798) | 0.143 (p=0.331) | 0.187 (p=0.203) |
| 3.30pm cortisol day 3 | 0.168 (p=0.269) | 0.381 (p=0.010)* | 0.362 (p=0.015)* | -0.108 (p=0.505) | -0.054 (p=0.725) | 0.230 (p=0.129) |
| Ratio am:pm cortisol day 3 | -0.002 (p=0.988) | 0.009 (p=0.955) | 0.159 (p=0.308) | 0.004 (p=0.983) | 0.159 (p=0.309) | 0.203 (p=0.191) |
| 9am cortisol day 5 | 0.297 (p=0.012) | 0.139 (p=0.251) | 0.232 (p=0.054) | -0.066 (p=0.609) | 0.012 (p=0.920) | -0.085 (p=0.486) |
| 3.30pm cortisol day 5 | 0.392 (p=0.001)** | 0.075 (p=0.547) | 0.237 (p=0.055) | 0.085 (p=0.513) | 0.02 (p=0.874) | -0.098 (p=0.432) |
| Ratio am:pm cortisol day 5 | -0.087 (p=0.493) | -0.091 (p=0.477) | -0.091 (p=0.472) | -0.017 (p=0.898) | 0.046 (p=0.720) | 0.062 (p=0.626) |
| 9am cortisol day 7 | -0.043 (p=0.762) | -0.170 (p=0.229) | -0.024 (p=0.868) | 0.024 (p=0.878) | -0.174 (p=0.216) | -0.036 (p=0.797) |
| 3.30pm cortisol day 7 | 0.139 (p=0.351) | -0.050 (p=0.737) | 0.243 (p=0.099) | -0.150 (p=0.357) | -0.104 (p=0.486) | -0.248 (p=0.093) |
| Ratio am:pm cortisol day 7 | -0.144 (p=0.346) | -0.118 (p=0.439) | -0.164 (p=0.281) | 0.082 (p=0.623) | -0.197 (p=0.195) | 0.149 (p=0.329) |
| 9am cortisol day 14 | 0.338 (p=0.041) | -0.052 (p=0.459) | 0.303 (p=0.068) | 0.134 (p=0.459) | 0.072 (p=0.670) | 0.338 (p=0.041)* |
| 3.30pm cortisol day 14 | 0.537 (p=0.002) | 0.225 (p=0.232) | 0.419 (p=0.021)* | 0.173 (p=0.379) | 0.084 (p=0.658) | 0.064 (p=0.737) |
| Ratio am:pm cortisol day 14 | -0.006 (p=0.978) | -0.193 (p=0.336) | -0.012 (p=0.954) | 0.147 (p=0.482) | -0.144 (p=0.473) | 0.218 (p=0.275) |
| 9am cortisol day 28 | 0.049 (p=0.672) | 0.046 (p=0.691) | 0.038 (p=0.740) | -0.010 (p=0.936) | -0.074 (p=0.521) | -0.001 (p=0.996) |
| 3.30pm cortisol day 28 | 0.390 (p=0.001) | 0.233 (p=0.045)* | 0.357 (p=0.002)** | -0.048 (p=0.698) | 0.273 (p=0.019) | -0.182 (p=0.120) |
| Ratio am:pm cortisol day28 | -0.203 (p=0.105) | -0.213 (p=0.089) | -0.220 (p=0.078) | 0.008 (p=0.950) | -0.219 (p=0.08) | -0.11 (p=0.932) |
| 9am cortisol 4 months | 0.168 (p=0.181) | 0.122 (p=0.333) | -0.078 (p=0.536) | 0.154 (p=0.236) | -0.045 (p=0.721) | -0.123 (p=0.328) |
| 3.30pm cortisol 4 months | 0.072 (p=0.541) | 0.109 (p=0.356) | 0.113 (p=0.336) | 0.130 (p=0.291) | 0.307 (p=0.008)** | -0.140 (p=0.234) |
| Ratio am:pm cortisol 4 months | 0.033 (p=0.805) | -0.070 (p=0.600) | -0.165 (p=0.212) | -0.011 (p=0.937) | -0.251 (p=0.055) | -0.112 (p=0.400) |
| 9am cortisol 12 months | -0.250 (p=0.04) | 0.065 (p=0.598) | -0.252 (p=0.038)* | -0.053 (p=0.675) | 0.053 (p=0.669) | -0.094 (p=0.445) |
| 3.30pm<br>cortisol 12<br>months | 0.094 (p=0.447) | 0.068 (p=0.587) | -0.051 (p=0.681) | 0.103 (p=0.422) | 0.207 (p=0.093) | -0.105 (p=0.399) |
| Ratio am:pm<br>cortisol 12<br>months | -0.169 (p=0.185) | -0.012 (p=0.925) | -0.058 (p=0.651) | -0.138 (p=0.297) | -0.129 (p=0.313) | -0.308 (p=0.014)* |

\*significant at the 0.05 level
\*\*significant at the 0.01 level
\*\*\*significant at the <0.001 level

## Discussion

This study is the largest, and most detailed, study to date, to examine the associations between cortisol levels and delirium in the first 12 months after stroke. In bivariate analyses, higher afternoon cortisol levels, but not morning cortisol, were associated with delirium and its severity in the first 14 days after stroke. However, our multivariable analyses showed that only age and NIHSS score were independently associated with delirium.

It is possible that we missed a small effect size because we did not recruit our target of 120 patients and the incidence of delirium was lower than predicted. Also it’s possible that we might have missed short episodes of delirium between our assessments, particularly if the delirium was hypoactive. On the other hand, it’s plausible that the incidence of delirium in this study was lower than in previous studies, because of improvements in stroke unit care.

A larger study with greater power would be needed to establish whether cortisol truly has an independent effect on delirium status, or whether it is simply a marker of other factors, such as stroke severity. It must be noted that large studies in this patient population are difficult to do, because participants are often very unwell following stroke and yet must be assessed for delirium and have cortisol levels measured regularly. Proxy consent is often required (either because of the severity of stroke or because of pre-existing dementia) and this, along with the fact that a long-term follow-up period is desirable (because delirium can persist) make recruitment challenging. These difficulties are reflected in the small sample sizes in previous studies. Meta-analysis would also be challenging with the studies currently available, as methodologies employed vary considerably from study to study, particularly in the method of cortisol measurement.

Stroke severity (as measured by the NIHSS score) and age were both independently associated with delirium in this study. This is interesting, and supports the existing evidence with respect to predictors of delirium after stroke. A large study of delirium after stroke (n=527) found that stroke severity and age were independent predictors of delirium (although age only became an independent predictor if brain atrophy was left out of the model, presumably because the two may co-vary), and as previously described these factors both have plausible pathophysiological mechanisms which might explain their consistent association with delirium [26]. Age is associated with neurone loss, reduced blood flow to the brain, and reduced vascular density in the brain, all of which may increase susceptibility to delirium. Pathological processes in the brain such as those seen in vascular dementia or Alzheimer’s dementia are also more common in the ageing brain, and these also increase the risk of delirium [27]. Stroke severity is a complex variable, as the severity score may not always correlate with the actual size of the brain lesion, although in many cases it will. Furthermore, stroke severity is not necessarily a marker of overall illness severity, although again there is often a correlation between the two (and those with a large TACS are also more at risk of intercurrent illness, such as pneumonia). However, overall the stroke severity score is a marker of the direct brain insult and subsequent brain tissue damage and loss [28], all of which are likely to increase the risk of delirium.

Ahmed and colleagues [29] examined salivary cortisol levels after stroke (rather than plasma or urinary levels). It found a mean salivary cortisol level (at admission) of 18.4nmol/L in the morning and 6.7nmol/L in the afternoon. In the current study, we found a mean morning level of 25.9nmol/L and a mean afternoon level of 17.7 nmol/L during the first week after stroke. The reason for the slightly higher levels in this study are not clear. It may partly be accounted for by the fact that collection devices and assays used differed, however assays in this study were calibrated to industry standards. Stroke severity is known to influence cortisol levels in a bidirectional fashion, in that those with very severe strokes in ICU have been shown to have low cortisol levels and lose the diurnal circadian rhythm, whereas more generally the more severe the stroke, the higher the cortisol level is [8]. The mean NIHSS score in the Ahmed study was 7, and the median NIHSS in this study was 5, indicating similar stroke severity. The Ahmed study was smaller (n=58) and the study participants were younger (mean age of 66, compared with 77 in this study), and although delirium wasn’t reported, it’s incidence is likely to have been lower, because participants were younger (and therefore less likely to have underlying risk factors for delirium such as pre-existing dementia).

There are some limitations to this study. Participants were recruited as soon after stroke as possible, however because the time of stroke onset was taken to be when the participant was last seen well, there were delays between this time and first assessment. There were also delays when proxy consent was required, as proxies, understandably, often required 24 hours or more to consider the study before granting consent. These factors meant that some participants were not assessed, nor did they have salivary cortisol measured until day 5 after stroke onset. Participants were not assessed every day following recruitment, rather they were assessed on alternate days. This methodology was used to reduce the burden of assessments for participants, however the chart-CAM and clinician and informant information was used to provide information about the days when formal assessments weren’t scheduled. It is possible that brief episodes of delirium were missed, although this could also be the case if participants were assessed daily. There was selection bias in the study, as proxies were more likely to decline consent compared with potential participants themselves, resulting in a bias towards selection of those with capacity at recruitment. Finally, as discussed above, the sample size was small, although this remains the largest study to date of cortisol and delirium after stroke.

In summary, we did not find evidence of an independent association between salivary cortisol levels after stroke and the development of delirium. Because the study was slightly underpowered, we cannot exclude for certain a relationship, but if such a relationship does exist, then it is likely to be small, and may not be of clinical relevance. Our data could be used to inform the design of future larger definitive studies. The focus of future studies should be on alternative mechanisms for delirium after stroke.

## Data Availability

Additional data may be available on request

## Acknowledgements

We are grateful to Dr Roanna Hall for helping to train Dr Barugh in assessment of delirium, and for advice from Professor John O’Brien, Professor John Starr, Professor Martin Dennis and Dr John McManus, the staff at the Clinical Research Facility at the Royal Infirmary of Edinburgh for their assistance processing samples, and the staff at the Acute Stroke Unit at the Royal Infirmary of Edinburgh for assistance recruiting patients.

Dr Barugh was funded by a Research Training Fellowship from the Dunhill Medical Trust. Alasdair MacLullich, Amanda Barugh, Susan Shenkin and Michael Allerhand were members of the Centre for Cognitive Ageing and Cognitive Epidemiology funded by BBSCR, ESRC, MRC as part of the Lifelong Health and Well-being Initiative.

We are grateful to Scotland A research ethics committee for approving the study.

---

**Figure 1.**
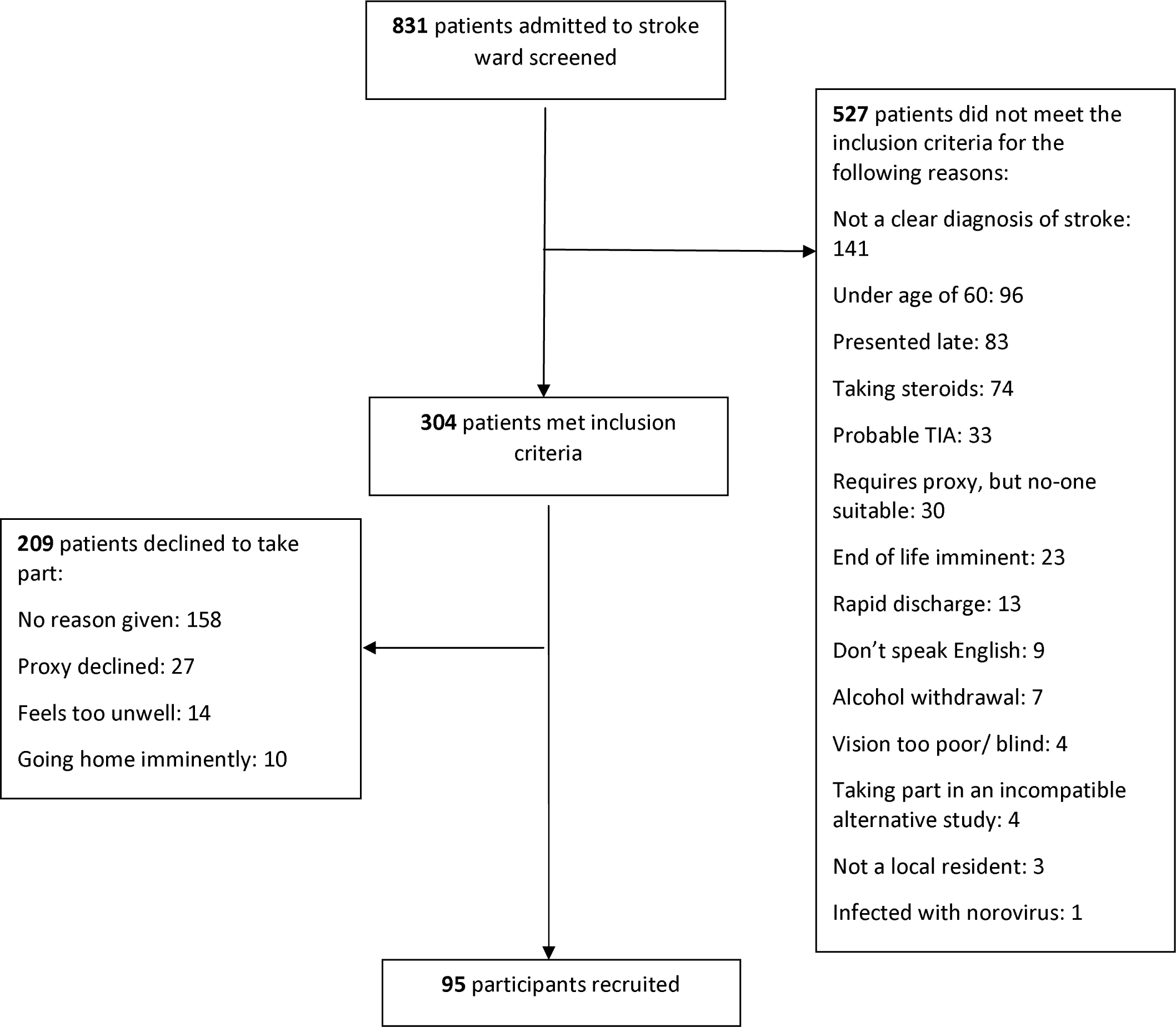
Flow diagram for recruitment

## References

[1] Stokholm J, Steenholt JV, Csilag C, Kjær TW, Christensen T. Delirium Assessment in Acute Stroke: A Systematic Review and Meta-Analysis of Incidence, Assessment Tools, and Assessment Frequencies. J Cent Nerv Syst Dis. 2019 Dec 30;11:1179573519897083. doi: 10.1177/1179573519897083. PMID: 31908562; PMCID: PMC6937530.

[2] Shaw R, Drozdowska B, Taylor-Rowan M, Elliot E, Cuthbertson G, Stott DJ, Quinn DT. Delirium in an Acute Stroke Setting, Occurrence, and Risk Factors Stroke. 2019;50:3265–3268

[3] Rollo E, Brunetti V, Scala I, Callea A, Marotta J, Vollono C, Frisullo G, Broccolini A, Calabresi P, Della Marca G. Impact of delirium on the outcome of stroke: a prospective, observational, cohort study. J Neurol. 2022 Dec;269(12):6467–6475. doi: 10.1007/s00415-022-11309-2. Epub 2022 Aug 9. PMID: 35945396; PMCID: PMC9618551.

[4] Pasińska P, Wilk A, Kowalska K, Szyper-Maciejowska A, Klimkowicz-Mrowiec A. The long-term prognosis of patients with delirium in the acute phase of stroke: PRospective Observational POLIsh Study (PROPOLIS). J Neurol. 2019; 266(11):2710–2717

[5] Richardson SJ, Davis DHJ, Stephan BCM, Robinson L, Brayne C, Barnes LE, Taylor J-P, Parker SG, Allan LM, Recurrent delirium over 12 months predicts dementia: results of the Delirium and Cognitive Impact in Dementia (DECIDE) study, Age and Ageing, 2021; 50, 914–920

[6] Cao Y, Song Y, Ding Y, Ni J, Zhu B, Shen J, Miao L. The role of hormones in the pathogenesis and treatment mechanisms of delirium in ICU: The past, the present, and the future. J Steroid Biochem Mol Biol. 2023 Oct;233:106356. doi: 10.1016/j.jsbmb.2023.106356. Epub 2023 Jun 28. PMID: 37385414

[7] MacLullich AMJ, Ferguson KJ, Miller T, De Rooij SEJA, Cunningham C. Unravelling the pathophysiology of delirium: A focus on the role of aberrant stress responses. Journal of Psychosomatic Research, 2008;65 (3), 229–238.

[8] Barugh AJ, Gray P, Shenkin SD, MacLullich AM, Mead GE Cortisol levels and the severity and outcomes of acute stroke: a systematic review. J Neurol, 2014;261, 533–45.

[9] Tene O, Hallevi H, Korczyn AD, Shopin L, Molad J, Kirschbaum C, Bornstein NM, Shenhar-Tsarfaty S, Kliper E, Auriel E, Usher S, Stalder T, Ben Assayag E. The Price of Stress: High Bedtime Salivary Cortisol Levels Are Associated with Brain Atrophy and Cognitive Decline in Stroke Survivors. Results from the TABASCO Prospective Cohort Study. J Alzheimers Dis. 2018;65(4):1365–1375. doi: 10.3233/JAD-180486. PMID: 30149451.

[10] Gustafson YOT, Asplund K, Hagg E. Acute confusional state (delirium) soon after stroke is associated with hypercortisolism. Cerebrovasc Dis, 1993; 3, 33–38.

[11] Fassbender K, Schmidt R, Mossner R, Daffertshofer M, Hennerici M Pattern of activation of the hypothalamic-pituitary-adrenal axis in acute stroke: Relation to acute confusional state, extent of brain damage, and clinical outcome. Stroke, 1994; 25 (6), 1105–1108.

[12] Marklund N, Peltonen M, Nilsson TK, Olsson T. Low and high circulating cortisol levels predict mortality and cognitive dysfunction early after stroke. Journal of Internal Medicine, 2004; 256 (1), 15–21.

[13] Olsson T. 1990. Urinary free cortisol excretion shortly after ischaemic stroke. Journal of Internal Medicine, 1990g; 228(2), 177-181.

[14] McGurn B, Starr JM, Topfer JA, Pattie A, Whiteman MC, Lemmon HA, Whalley LJ, Deary IJ. Pronunciation of irregular words is preserved in dementia, validating premorbid IQ estimation. Neurology 2004, 62, 1184–6.

[15] Jorm A A short form of the informant questionnaire on cognitive decline in the elderly (iqcode): Development and cross-validation. Psychol Med, 1995; 25, 437.

[16] Chikakuta AAA. Audit in intensive care. The APACHE II classification of severity of disease. Ulster Med J, 1990;59, 161–167.

[17] Ely E, Inouye SK, Bernard GR, Gordon S, Francis J, May L, Truman B, Speroff T, Gautam, S, Margolin, R, Hart, RP, Dittus, R 2001. Delirium in mechanically ventilated patients validity and reliability of the confusion assessment method for the intensive care unit (cam-icu). JAMA - Journal of the American Medical Association, 286, 2703–2710.

[18] Sessler CGM, Grap MJ, Brophy GM, O’Neal PV, Keane KA, Tesoro EP, Elswick RK. The Richmond Agitation-Sedation Scale: validity and reliability in adult intensive care unit patients. Am J Respir Crit Care Med. 2002, 166, 1338–1344.

[19] Trzepacz PMD, Torres R, Kanary K, Norton J, Jimerson N 2001. Validation of the delirium rating scale-revised-98. Comparison with the delirium rating scale and the cognitive test for delirium. 2001; J Neuropsychiatrt Clin Neurosci, 13, 229-242.

[20] Tieges Z, Mcgrath A, Hall RJ, Maclullich AM. Abnormal level of arousal as a predictor of delirium and inattention: an exploratory study. Am J Geriatr Psychiatry, 2013; 21, 1244–1253.

[21] Inouye SK, Leo-Summers L, Zhang Y, Bogardus ST, Jr., Leslie DL, Agostini JV. A chart-based method for identification of delirium: validation compared with interviewer ratings using the confusion assessment method. J Am Geriatr Soc, 2005; 53, 312–8.

[22] Brown LJ, Fordyce C, Zaghdani H, Starr JM, MacLullich AM. Detecting deficits of sustained visual attention in delirium. J Neurol Neurosurg Psychiatry. 2011 Dec;82(12):1334–40. doi: 10.1136/jnnp.2010.208827. Epub 2010 Jun 28. PMID 20587493.

[23] Nasreddine ZS, Phillips NA, Bedirian V, Charbonneau S, Whitehead V, Collin I, Cummings, JL, Chertkow H. The Montreal Cognitive Assessment, MoCA: A Brief Screening Tool For Mild Cognitive Impairment. JAGS, 2005; 53, 695-699.

[24] Pendlebury ST, Cuthbertson FC, Welch JV, Mehta Z, Rothwell PM. Underestimation of Cognitive Impairment by Mini-Mental State Examination Versus the Montreal Cognitive Assessment in Patients With Transient Ischemic Attack and Stroke. Stroke, 2010 41, 1290–1293.

[25] Lincoln NB, Gladman JR The Extended Activities of Daily Living scale: a further validation. Disabil Rehabil, 1992; 14, 41–3.

[26] Oldenbeuving AW, de Kort PL, Jansen BP, Algra A, Kappelle LJ, Roks G. Delirium in the acute phase after stroke: incidence, risk factors, and outcome. Neurology. 2011 Mar 15;76(11):993–9. doi: 10.1212/WNL.0b013e318210411f. Epub 2011 Feb 9. PMID: 21307355.

[27] Maldonado, J. R. 2013. Neuropathogenesis of Delirium: Review of current Etiologic Theories and Common Pathways. Am J Geriatr Psychiatry, 21, 1190–1222.

[28] Mitsias PD, Jacobs MA, Hammoud R, Pasnoor M, Santhakumar S, Papamitsakis NI, Soltanian-Zadeh H, Lu M, Chopp M, Patel SC. Multiparametric MRI ISODATA ischemic lesion analysis: correlation with the clinical neurological deficit and single-parameter MRI techniques. Stroke, 2002 33, 2839–44.

[29] Ahmed N, De La Torre B, Wahlgren NG. Salivary cortisol, a biological marker of stress, is positively associated with 24-hour systolic blood pressure in patients with acute ischaemic stroke. Cerebrovascular Diseases, 2004;18 (3), 206–213.

